# Characterizing emergency clinician engagement with social drivers of health data among patients with opioid use disorder

**DOI:** 10.64898/2025.12.15.25342249

**Authors:** Melanie F. Molina, Samuel D. Pimentel, Cynthia Fenton, Julia Adler-Milstein, Laura M. Gottlieb

**Affiliations:** Department of Emergency Medicine, University of California, San Francisco, California, USA; Department of Statistics, University of California, Berkeley, Berkeley, California, USA; Department of Medicine, University of California, San Francisco, San Francisco, California, USA; Center for Clinical Informatics & Improvement Research, University of California, San Francisco, San Francisco, California, USA; Social Interventions Research and Evaluation Network, University of California, San Francisco, San Francisco, California, USA; Department of Family and Community Medicine, University of California, San Francisco, San Francisco, California, USA

**Author notes:** **Address manuscript correspondence to:** Melanie F. Molina, MD, MAS, UCSF Department of Emergency Medicine, Room L-126, Box 0209, 505 Parnassus Avenue, San Francisco, CA 94143.

**Keywords:** social drivers of health, electronic health records, emergency medicine, opioid use disorder, documentation, review, medication for opioid use disorder

## Abstract

**Objectives:** To characterize emergency department (ED) clinician engagement with electronic health record (EHR)-based social drivers of health (SDOH) data; test whether engagement differs in encounters with opioid use disorder (OUD); and, among OUD encounters, assess whether engagement is associated with medications for OUD (MOUD).

**Materials and Methods:** We conducted a cross-sectional study of adult ED encounters (January 2023–October 2024). OUD encounters, identified with a structured phenotype, were matched (1:2) to non-OUD encounters. Audit logs captured clinician engagement with structured SDOH questions (“SDOH Wheel”), ICD-10 Z codes in the Problem List, Social History free text, and social work notes. Engagement was any SDOH documentation or review of preexisting SDOH data during the encounter. Logistic regression estimated associations.

**Results:** Among 17,103 encounters (5,701 OUD; 11,402 non-OUD), clinician SDOH documentation was rare (<1%). Clinicians most often reviewed Z codes (610/620; 98.4%), followed by the SDOH Wheel (1,103/3,953; 27.9%), social work notes (1,711/10,670; 16.0%), and Social History free text (232/6,942; 3.3%). Engagement occurred in 19.5% of encounters and was higher with OUD (26.6% vs 16.0%; OR 1.91, 95% CI 1.77–2.07). Among OUD encounters, engagement was not associated with MOUD (OR 1.11, 95% CI 0.84–1.47); however, MOUD treatment varied by race and ethnicity, reflecting persistent disparities.

**Discussion:** ED clinicians infrequently documented but did review structured, accessible SDOH data. Engagement increased in OUD encounters yet neither predicted MOUD nor mitigated racial and ethnic treatment disparities. EHR interfaces that surface SDOH data coupled with targeted decision supports might influence equitable, time-sensitive ED care.

## BACKGROUND AND SIGNIFICANCE

Over the last decade, electronic health record (EHR)-based social drivers of health (SDOH) documentation tools have rapidly proliferated,^1^ driven in part by reimbursement and accreditation policies that incentivize identifying and addressing patient-level adverse SDOH, or social risks, in clinical settings.^2–4^ Structured EHR-based SDOH data are intended to improve outcomes by informing clinical decisions around treatment and resource linkages. Yet relatively little research has examined clinician engagement with available SDOH data or how engagement with SDOH data affects care, in part, because of difficulties assessing when and how clinicians use EHR-based information. The capacity to leverage audit log data—systematic records of user activities—however, is likely to help researchers overcome these barriers. In fact, one new study used audit log data to examine both clinician documentation of SDOH data and review of prior SDOH data^1^ and found that both measures were associated with lower risk of hospital readmission.^5^ Using audit log data and other EHR-facilitated data tools to better understand the impacts of SDOH data documentation is especially critical in the context of recent federal proposals to decrease SDOH-related healthcare incentives.^6^

The emergency department (ED) often serves as the primary healthcare access point for vulnerable populations,^7^ including those with a high prevalence of adverse SDOH,^8^ making it an important place for social risk screening and interventions. To explore how ED clinicians engage with EHR-based SDOH data and whether engagement impacts clinical treatment decisions, we selected opioid use disorder (OUD) as a condition-specific use case. OUD afflicts approximately 2.7 million people in the US,^9^ and evidence-based medications for OUD (MOUD), such as buprenorphine and methadone,^10^ are increasingly administered in the ED.^11^ Since adverse SDOH are common among patients experiencing OUD and affect treatment outcomes,^12,13^ one might expect that ED clinicians would engage more with SDOH data during encounters with patients diagnosed with OUD compared to those without and then use SDOH information to guide treatment decisions.

## OBJECTIVES

In this study, we addressed three novel research questions using ED visit EHR data. First, we characterized ED clinician engagement with EHR-based SDOH data by assessing overall documentation and review.

Second, we examined whether ED clinician engagement with SDOH data was associated with OUD—a condition highly impacted by SDOH.^14^ Third, we determined whether engagement with SDOH data in patients with OUD was associated with downstream clinical action by assessing the association between engagement with SDOH data and MOUD treatment, which could inform the design of future interventions to optimize ED-based OUD care delivery.

## METHODS

### Design, setting, and study sample

We conducted a cross-sectional study of adult ED visit encounters at the University of California, San Francisco (UCSF) Medical Center from January 1, 2023 through October 17, 2024—five years after the systemwide rollout of new SDOH documentation tools. UCSF Medical Center is a quaternary academic center and houses UCSF Health’s only adult ED, which serves over 40,000 patients annually and has used the Epic EHR since 2012. We included encounters for adults (≥18 years) meeting an EHR-derived phenotype for OUD, plus propensity score-matched encounters without OUD. The OUD phenotype was adapted from a previously validated computable structured phenotype^15^ and broadened to improve sensitivity and align with our EHR, as formal OUD documentation is often incomplete due to stigma and variable clinical practices. Our site-adapted OUD phenotype consisted of any of the following criteria, applied to the entire hospital encounter unless otherwise specified: (1) one or more International Classification of Diseases, Tenth Revision (ICD-10) codes indicating OUD, withdrawal, or overdose in encounter (discharge) diagnoses, problem lists, or past medical history; (2) active prescriptions for buprenorphine or methadone preceding the index ED visit encounter; (3) positive urine toxicology results for methadone, fentanyl, or heroin; (4) addiction care team consultation notes with reason for consult containing “heroin”, “opioid”, “opiate”, “Narcan”, and/or “fentanyl”; (5) chief complaint containing “heroin”, “opioid”, “opiate”, and/or “OUD”, “fentanyl” (excluding use together with phrases suggesting therapeutic use or emergency medicine services-only administration), “fentanyl use”, and/or mapping to an ICD-10 code for OUD; (6) reason for visit free text containing the same key words as in (5); or (7) ED clinician notes (including triage nursing notes) containing “OUD”, “opioid use disorder”, “opioid use d/o”, “COWS” (Clinical Opiate Withdrawal Scale), “fentanyl”, and/or “heroin”.

To compare engagement with SDOH data among patients with versus without OUD, we designed a matched non-OUD comparison group. We used propensity scores to match each OUD patient to two patients without OUD. We estimated propensity scores using logistic regression on all ED encounters during the study period using age, ED arrival time, language, interpreter need, marital status, race and ethnicity, sex assigned at birth, gender identity, sexual orientation, insurance type, and zip code (coarsened by grouping zip codes fewer than 100 visits). To ensure covariate balance, we imposed a fine balance constraint on the interaction of race and ethnicity, marital status, sex assigned at birth, sexual orientation, and an indicator for rare zip codes.^16,17^ The UCSF Institutional Review Board approved this study, and we followed the Strengthening the Reporting of Observational Studies in Epidemiology (STROBE) reporting guidelines.

### Selected EHR-based SDOH data field types

ED clinicians can interface with four distinct types of SDOH data within the EHR. These include two structured data sources: (1) Implemented in Epic in 2018, structured social history questions (i.e., the “Epic SDOH Wheel”) capture information on housing, transportation, and financial status.^1^ These fields offer responses ranging from “Never true” to “Often true” with a “Declined to Answer” option. UCSF Health encourages clinical staff to complete SDOH data fields in both inpatient and outpatient settings, though there is no official ED-based documentation workflow. Nevertheless, the SDOH data fields are easily visible to ED clinicians in Epic’s storyboard; (2) Structured social risk ICD-10 Z codes enable clinicians to document adverse SDOH as part of the past medical history and can populated in any clinical setting. ED clinicians can view and edit these codes as part of the patient’s Problem List directly from the ED navigation screen without opening a patient’s chart. Two unstructured SDOH data sources include: (3) “Social History” free text, which is a subsection of the Patient History; and (4) Social work free text notes, which are only available for patients evaluated by social work.

### Definitions and measures

Using audit log data, we assessed whether one or more of these SDOH data types were documented or reviewed by at least one ED clinician during the ED encounter (time-bound from ED arrival to disposition). Any documentation or review event occurring after the ED encounter was excluded. “Clinicians” included attending physicians, resident physicians, and advanced practice providers. Given that multiple handoffs between clinicians occur during a single ED visit, we only included clinicians assigned to the care team for more than 10 minutes or if they authored/cosigned a note during the encounter from the EHR’s ED context.

An ED clinician documentation event occurred if SDOH data (whether adverse or not) were entered into any of the following data fields: (1) the SDOH Wheel, (2) the Problem List (SDOH Z codes, adverse by definition), (3) the Social History subsection of the Patient History (free text). Review events occurred if the ED clinician: (1) hovered over or clicked any section of the SDOH Wheel; (2) viewed the Problem List from the ED navigation screen (via a hover pop-up) or marked it as reviewed; (3) marked the Social History free text field as reviewed; or (4) opened a social work note (data review only). We defined two types of review events. *Field review* occurred when a clinician reviewed a data field regardless of whether it contained SDOH data (possible for the SDOH Wheel, Social History free text, and Problem List; social work notes have no blank data field). *Data review* occurred when the clinician reviewed a data field populated with preexisting SDOH data. We assessed both field and data review events. However, we defined ED clinician “engagement” with SDOH data as one or more instances in which the clinician documented or reviewed any SDOH data (i.e., data review) during the ED encounter.

For the structured data types (i.e., SDOH Wheel and Z codes), we categorized the SDOH data into financial, housing, transportation, food, and safety domains. For the SDOH Wheel, in addition to the simple presence or absence of SDOH data, we were able to further specify whether SDOH data were present and adverse (i.e., documented social risks). We therefore defined a sub-measure of engagement, “risk engagement,” as engagement with the SDOH Wheel when adverse SDOH data were present.

For each ED encounter, we also extracted patient age, gender, self-reported race and ethnicity, language, insurance class as well as MOUD treatment, defined as either an ED order to administer buprenorphine or methadone, or an ED-initiated outpatient prescription for buprenorphine (ED clinicians cannot prescribe methadone). Since patients could have multiple ED encounters during the study period, and our focus was on clinician behaviors, we derived our measures at the encounter level rather than the patient level. We counted documentation and review measures once per encounter, regardless of how many clinicians were involved.

### Outcomes

Our primary outcome was overall ED clinician engagement with SDOH data during the ED encounter. We then compared engagement with SDOH data between patients with and without OUD, and, among patients with OUD, examined the association between engagement and MOUD treatment. Finally, to understand whether previously documented social risks affected MOUD treatment, we also assessed whether risk engagement was associated with MOUD.

### Statistical analysis

We used descriptive statistics to summarize ED clinician documentation, field and data review, and engagement with each SDOH data field at the encounter level across all encounters (research question 1). Results were stratified by patient group (OUD versus non-OUD) and SDOH domain. We then used multivariable logistic regression to assess whether ED clinician engagement with SDOH data differed among encounters with patients experiencing OUD compared to matched controls (research question 2) and whether MOUD treatment differed among OUD encounters with engagement and/or risk engagement (research question 3). We adjusted for key patient demographic characteristics known or hypothesized to be associated with SDOH engagement and MOUD treatment, including age, race and ethnicity, sex, and insurance status.^5,18,19^

We conducted several sensitivity analyses. For the MOUD treatment model, we excluded encounters where patients met the OUD phenotype solely because they were already on MOUD to assess whether engagement with SDOH data was associated with MOUD initiation. For both the engagement and MOUD treatment models, we attempted mixed-effects analyses clustered by clinician, but they could not be fit due to clinicians being cross-classified across encounters. Therefore, to verify the robustness of our findings, we compared models with and without clinician fixed effects using likelihood-ratio tests. All statistical analyses were performed using R statistical software version 4.4.2 (R Project for Statistical Computing).

## RESULTS

### ED encounter characteristics

Of 70,880 ED encounters during the study period, 5,701 (33%) encounters met the OUD phenotype criteria and were matched with 11,402 (67%) non-OUD encounters, yielding 17,103 total study encounters. **Supplementary Appendix A** shows the frequency of each OUD phenotype criterion across OUD encounters. Among the analyzed encounters, there were 477 unique clinicians, with a median (IQR) of 13 (2-85) encounters per clinician. There were 10,468 unique patients; 74% of these patients had a single ED encounter, another 20% had 2-3 encounters, and the remaining 7% had 4 or more. 43.8% of encounters were with patients who were White, 26.4% with patients who were Black, and 12.0% with patients who were Hispanic/Latino/a/x; 60.8% were with patients who were male; 51.7% were with patients enrolled in Medicaid. MOUD treatment was present in 387 (2.3%) encounters, 362 (93.5%) of these meeting the OUD phenotype and 25 (6.5%) not (**Table 1**).

**Table 1.** Emergency Department Encounter Characteristics, by Opioid Use Disorder Status.

|  | Total Encounters<br>N (%) | OUD Phenotype Met<br>n (%) | OUD Phenotype Not Met<br>n (%) |
| --- | --- | --- | --- |
| **Overall** | 17103 (100%) | 5701 (33.3%) | 11402 (66.7%) |
| **Unique Patients<sup>a</sup>** | 10,468 (100%) | 2,757 (26.3%) | 8,042 (76.8%) |
| **Age, Mean (Std. Dev.)** | 51.1 (16.1) | 51.1 (16.0) | 51.1 (16.2) |
| **Race and Ethnicity** |  |  |  |
| White | 7487 (43.8%) | 2524 (44.3%) | 4963 (43.5%) |
| Asian | 1029 (6.0%) | 337 (5.9%) | 692 (6.1%) |
| Black or African American | 4510 (26.4%) | 1484 (26.0%) | 3026 (26.5%) |
| Hispanic or Latino/a/x | 2057 (12.0%) | 676 (11.9%) | 1381 (12.1%) |
| Multi-racial | 720 (4.2%) | 250 (4.4%) | 470 (4.1%) |
| Native American, Alaska Native, or Other Pacific Islander | 179 (1.0%) | 65 (1.1%) | 114 (1.0%) |
| Other<sup>b</sup> | 1121 (6.6%) | 365 (6.4%) | 756 (6.6%) |
| **Sex** |  |  |  |
| Male | 10403 (60.8%) | 3478 (61.0%) | 6925 (60.7%) |
| Female | 6618 (38.7%) | 2187 (38.4%) | 4431 (38.9%) |
| Other<sup>c</sup> | 82 (0.5%) | 36 (0.6%) | 46 (0.4%) |
| **Insurance** |  |  |  |
| Medicaid | 8850 (51.7%) | 2950 (51.7%) | 5900 (51.7%) |
| Medicare | 5658 (33.1%) | 1886 (33.1%) | 3772 (33.1%) |
| Commercial | 1914 (11.2%) | 638 (11.2%) | 1276 (11.2%) |
| Other<sup>d</sup> | 681 (4.0%) | 227 (4.0%) | 454 (4.0%) |
| **MOUD Treatment** | 387 (2.3%) | 362 (6.3%) | 25 (0.2%) |

**Abbreviations:** OUD, opioid use disorder; MOUD, medication for opioid use disorder
<sup>a</sup>331 unique patients had OUD status that changed over time.
<sup>b</sup>Included “Other”, “Unknown/Declined”, “Southwest Asian and North African”
<sup>c</sup>Included “Choose not to disclose”, “Declines to answer”, “Something else”, “Unknown”
<sup>d</sup>Included “Other” and “Worker’s Comp”

### ED clinician documentation and review

The percentage of total encounters for which ED clinicians *documented* SDOH data across any data field (excluding social work notes since these were documented by social workers whose job includes documenting social history) was very low (<1%). When we examined review of the SDOH data fields, regardless of whether they contained SDOH data (i.e., field review), clinicians most often reviewed the Problem List data field (16,607/17,103; 97.1%), followed by the SDOH Wheel data fields (3,723/17,103; 21.8%), and the Social History free text data field (621/17,1013; 3.6%); there was no specific data field to review for social work notes (i.e., if no social work note existed, we could not assess whether the clinician looked for it) (**Figure 1**).

**Figure 1.**
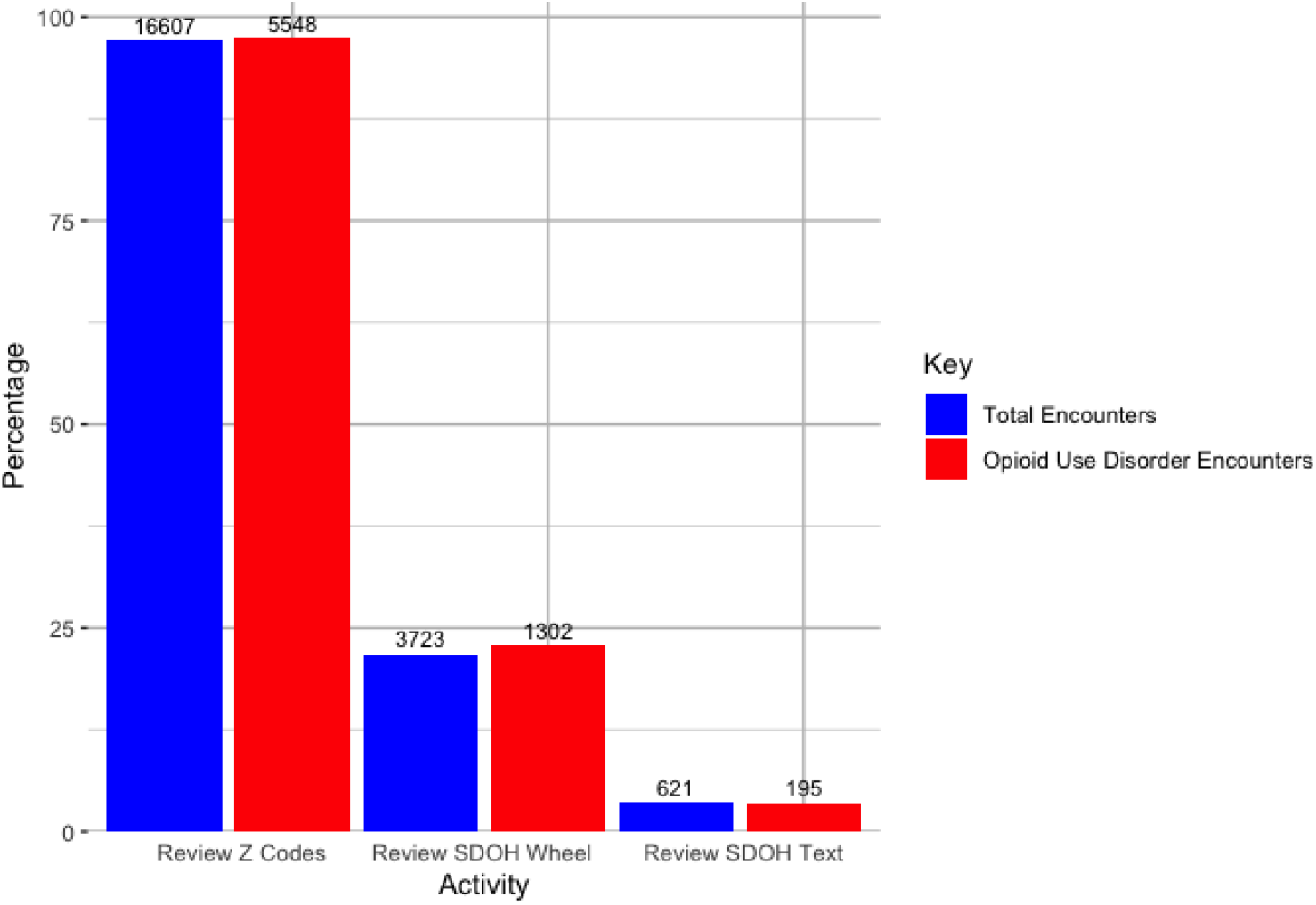
Emergency Department Clinician SDOH Data Field Review, by Type. **Abbreviations:** SDOH, social drivers of health

When we examined ED clinician review behavior of data fields with preexisting SDOH data (i.e., data review), we saw similar trends, though clinicians were more likely to review SDOH data fields when SDOH data were already documented (**Figure 2**). Clinicians most often reviewed Z codes (610/620; 98.4%), followed by the SDOH Wheel data (1,103/3,953; 27.9%), social work notes (1,711/10,670; 16.0%), and Social History free text data (232/6,942; 3.3%). Summing across all documentation and data review activities, engagement with SDOH data was present in 19.5% (3,343/17,103) of all encounters. Engagement was greater in encounters with patients experiencing OUD (26.6%; 1,515/5,701) compared to non-OUD (16.0%; 1,828/11,402).

**Figure 2.**
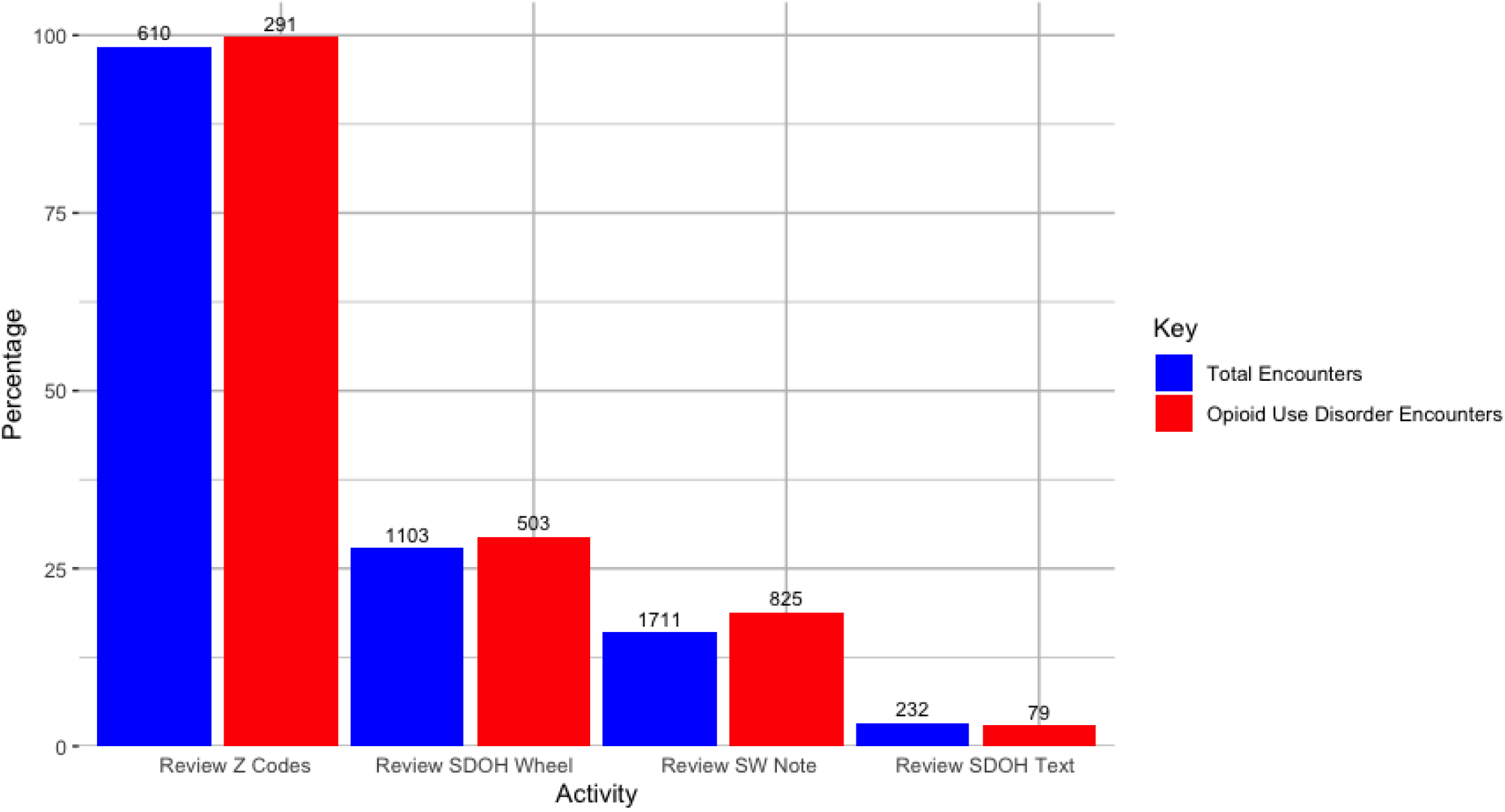
Emergency Department Clinician SDOH Data Review, by Type. **Abbreviations:** SDOH, social drivers of health; SW, social work

Regardless of whether reviewed or not, preexisting SDOH Wheel data were available for 3,953 encounters. This included 3,143 encounters containing data related to patient finances, 1,159 related to housing, 3,273 related to transportation, and 3,276 related to food. The SDOH Wheel most often contained housing data (38.9%), followed by food (29.6%), finance (29.2%), and transportation (28.3%). The most common preexisting Z codes related to housing (405), followed by food (182), finance (82), and safety (28).

Of the 3,953 encounters with preexisting SDOH Wheel data, 3,301 (83.5%) had documented adverse SDOH. Among these, provider risk engagement (i.e., engagement with the SDOH Wheel when adverse SDOH were already documented) occurred in 29.5% (975/3,301) of encounters and was also higher in encounters with patients experiencing OUD: 31.1% (466/1,496) versus 28.2% (509/1,805).

### Associations between clinician engagement with SDOH data and MOUD treatment

After adjusting for age, race and ethnicity, sex, and insurance status, we found that clinicians were more likely to engage with SDOH data for patients with OUD (OR 1.91, 95% CI 1.77-2.07), and for Black/African American (OR 1.20, 95% CI 1.09-1.31), muti-racial (OR 1.35, 95% CI 1.13-1.63), Native American, Alaska Native, or Other Pacific Islander (OR 2.28, 95% CI 1.65-3.14), female (OR 1.19, 95% CI 1.10-1.28), and Medicare (OR 1.25, 95% CI 1.13-1.38) patients. They were less likely to engage with SDOH data for patients with Asian (OR 0.83, 95% CI 0.70-0.99) and other race (OR 0.57, 95% CI 0.46-0.70), and for those with commercial (OR 0.69, 95% CI 0.60-0.80) and other insurance (OR 0.27, 95% CI 0.19-0.38). Overall, engagement with SDOH data was not significantly associated with MOUD (OR 1.11, 95% CI 0.84–1.47). However, in stratified analyses, engaging with adverse SDOH was associated with lower MOUD treatment (OR 0.87, 95% CI 0.54–1.42) and engaging with non-adverse SDOH with higher MOUD treatment (OR 1.90, 95% CI 0.65–5.57), though neither was significant. MOUD treatment was less likely for Asian (OR 0.45, 95% CI 0.24-0.83), Black/African American (OR 0.71, 95% CI 0.54-0.93), and Hispanic/Latino/a/x (OR 0.48, 95% CI 0.32-0.72) patients (**Table 2**). Results were similar after removing encounters where patients met the OUD phenotype only because they were already on MOUD (5,372 encounters, or 94.2% of total OUD encounters). In this analysis, patients with commercial insurance were also less likely to receive MOUD (OR 0.60, 95% CI 0.38-0.92).

**Table 2.** Association of Patient Characteristics with Clinician Engagement and MOUD Treatment.

|  | Engagement<br>OR (95% CI) | MOUD Treatment<br>OR (95% CI) |
| --- | --- | --- |
| **Age** | 1.00 (1.00-1.01) | 0.99 (0.98-1.00) |
| **Race and Ethnicity** |  |  |
| White | Reference | Reference |
| Asian | 0.83 (0.70-0.99) | 0.45 (0.24-0.83) |
| Black or African American | 1.20 (1.09-1.31) | 0.71 (0.54-0.93) |
| Hispanic or Latino/a/x | 0.95 (0.83-1.08) | 0.48 (0.32-0.72) |
| Multi-racial | 1.36 (1.13-1.63) | 0.71 (0.40-1.25) |
| Native American, Alaska Native or Other Pacific Islander | 2.28 (1.65-3.14) | 0.76 (0.27-2.16) |
| Other<sup>a</sup> | 0.57 (0.46-0.70) | 1.18 (0.80-1.76) |
| **Sex** |  |  |
| Male | Reference | Reference |
| Female | 1.19 (1.10-1.28) | 0.92 (0.73-1.16) |
| Other<sup>b</sup> | 1.26 (0.74-2.16) | 0 (0-2.61<sub>E198</sub>)<sup>d</sup> |
| **Insurance** |  |  |
| Medicaid | Reference | Reference |
| Medicare | 1.25 (1.13-1.38) | 0.76 (0.56-1.03) |
| Commercial | 0.69 (0.60-0.80) | 0.69 (0.47-1.01) |
| Other<sup>c</sup> | 0.27 (0.19-0.38) | 0.87 (0.52-1.45) |
| **Encounter Phenotype** |  |  |
| Non-Opioid Use Disorder | Reference | - |
| Opioid Use Disorder | 1.91 (1.77-2.07) | - |
| **Engagement** | - | 1.11 (0.84-1.47) |
| **Risk Engagement** |  |  |
| SDOH Absent | - | Reference |
| SDOH Present and Non-Adverse | - | 1.90 (0.65-5.57) |
| SDOH Present and Adverse | - | 0.87 (0.54-1.42) |

**Abbreviations:** MOUD, medications for opioid use disorder; OR, odds ratio; SDOH, social drivers of health
<sup>a</sup>Includes “Other”, “Unknown/Declined”, “Southwest Asian and North African”
<sup>b</sup>Includes “Choose not to disclose”, “Declines to answer”, “Something else”, “Unknown”
<sup>c</sup>Includes “Other” and “Worker’s Comp”
<sup>d</sup>Confidence interval uninformative due to no MOUD treatment events for individuals in this category

Adding clinician fixed effects to the models produced similar results. Engagement patterns persisted, but the lower engagement for Asian patients was no longer statistically significant (OR 0.83; 95% CI 0.69–1.00). Seventy-four (15.5%) of the 477 fixed effects were significant in the engagement model. Medicare patients were less likely to receive MOUD (OR 0.68; 95% CI 0.49–0.96), while MOUD treatment disparities persisted for Asian (OR 0.47; 0.24–0.95), Black/African American (OR 0.64; 0.47–0.86), and Hispanic/Latino/a/x patients (OR 0.48; 0.30–0.75). Neither engagement (OR 0.87; 95% CI 0.63–1.21) nor risk engagement (OR 0.80; 0.45–1.41) was associated with MOUD treatment. In the MOUD model, only 6.2% of the 387 clinician fixed effects were significant—close to the 5% expected by chance—and a likelihood-ratio test showed that adding fixed effects did not improve model fit.

## DISCUSSION

In this cross-sectional study of 17,103 emergency department (ED) encounters, ED clinicians engaged with EHR-based SDOH data in fewer than 20% of encounters, primarily through data review rather than documentation. Engagement was approximately 10% higher for encounters involving OUD than for other encounters (27%). While engagement was not significantly associated with MOUD treatment, clinicians were twice as likely to provide MOUD when engaging with non-adverse SDOH compared with adverse SDOH. Racial and ethnic disparities in MOUD treatment persisted regardless of SDOH data engagement. To our knowledge, this is the first study to use clinician audit log data to examine how SDOH information is reviewed and documented in ED settings.

Though other authors have shown that ED clinicians rarely document information about patients’ SDOH,^20^ in this study, we also found that ED clinicians rarely review available SDOH information. The low rates of review suggest clinicians may be missing opportunities to address adverse SDOH and better tailor care to patients’ social circumstances. For patients with OUD—whose social context often influences adherence to MOUD^14^— SDOH data were reviewed slightly more often, yet still in fewer than one-third of encounters. These results suggest that expanding SDOH screening and EHR documentation is unlikely to affect ED care unless coupled with other interventions. To translate screening into clinical care changes, SDOH information must be surfaced at the right time and place in ED workflows.

Furthermore, ease of information retrieval appears critical in the time- and resource-constrained ED environment. Prior studies using similar engagement measures to examine the behaviors of inpatient clinical teams reported that clinicians reviewed social work notes in 70% of encounters, with Social History free text reviewed more frequently than the SDOH Wheel (14.4% vs 0.03%).^1^ In our study, clinicians preferentially reviewed structured elements such as Z codes and the SDOH Wheel. Although key study differences (e.g., prior work counted only “Mark as Reviewed” actions) complicate a direct comparison, the pattern suggests inpatient clinicians may rely more on unstructured notes, whereas time-pressed ED clinicians favor easily accessible structured data. The stark contrast in review rates between the Problem List (97.1%), which is visible directly from the ED trackboard, and other SDOH fields also supports this interpretation.

Although not statistically significant, the lower likelihood of MOUD treatment when clinicians viewed adverse versus non-adverse SDOH information in the EHR warrants further study. This pattern raises concern that SDOH documentation may inadvertently bias treatment decisions, which would not altogether be unexpected: prior work has shown that clinicians are hesitant to initiate MOUD when they anticipate barriers to follow-up.^21^ Clinicians reviewing adverse SDOH may expect poor care continuity, particularly given known challenges for patients with OUD facing adverse SDOH.^14^ The finding is also consistent with wide and persistent disparities in MOUD treatment practices for Asian, Black/African American, and Hispanic/Latino/a/x patients,^18,19^ who are more likely to face socioeconomic barriers to care.^22^ The risk of exacerbating bias in treatment practices might be mitigated by introducing policies that incentivize SDOH documentation alongside the provision of interventions that can mitigate the impact of adverse SDOH on adherence and follow-up. In other words, the presentation of adverse SDOH data may need to be coupled with actionable interventions or treatment adaptations. Clinical decision support tools—electronic health record (EHR)-embedded tools that guide clinical decision-making using order sets, reminders, and alerts—have been effective in influencing care, particularly when developed with user (e.g., clinician) input,^23–28^ and could be used to operationalize such support. Future research should explore these types of coupled interventions and evaluate their effect on equity in MOUD treatment.

As a single-site study conducted at an academic medical center, our findings may not be generalizable to other practice settings, particularly community EDs or those serving different patient populations. The socialized importance of SDOH at our institution likely led to higher clinician engagement than can be expected in other settings. While we were unable to cluster models by clinician, we verified robustness by adding clinician fixed effects, which did not meaningfully change our findings. Although EHR audit log data enabled us to measure documentation and review behaviors, the data do not provide a window into clinicians’ decision-making processes, or motivations for engaging with SDOH data. This limitation is particularly relevant to understanding the relationship between risk engagement and MOUD treatment, where qualitative research with clinicians might explore whether and how clinicians consciously factor adverse SDOH information into their treatment decisions.

Other limitations stem from our measures of SDOH data engagement. While we could track when clinicians accessed the problem list containing Z codes, we cannot guarantee they specifically reviewed the Z codes themselves versus other diagnoses. Additionally, because the SDOH Wheel was the only data field that distinguished between present, absent, and adverse SDOH data, we were likely insufficiently powered to detect significant associations between engagement and MOUD treatment. Nevertheless, our ability to classify the SDOH data within the SDOH Wheel provided a unique level of granularity that strengthened our analyses. Lastly, we were unable to categorize unstructured SDOH information from social work notes or the free text social history field into specific domains. However, these unstructured data fields represented a small proportion of overall engagement events in our sample, and categorizing SDOH domains was not the primary objective of this study.

## CONCLUSION

In summary, ED clinicians engaged with EHR-based SDOH information in fewer than 1 in 5 encounters, primarily through review of structured, easily accessible fields; new SDOH documentation was rare. Engagement with SDOH data was not significantly associated with MOUD treatment, yet we observed a concerning trend toward lower MOUD use when adverse SDOH were reviewed and higher MOUD use when non-adverse SDOH were reviewed. Racial and ethnic disparities in MOUD treatment persisted regardless of SDOH data engagement. These findings suggest that expanding SDOH screening and documentation alone is unlikely to change ED care. To translate SDOH data into equitable treatment, information must be surfaced at the right time and place in ED workflows and paired with actionable supports that address anticipated barriers (e.g., follow-up and adherence). Clinical decision support tools might be leveraged to operationalize such interventions. Future work should explore such interventions, testing their impact on improving treatment equity, as well as examine causal links between SDOH engagement and treatment decisions.

## Data Availability

Data may be made available upon request at the discretion of the authors and in compliance with institutional policies.

## AUTHOR CONTRIBUTIONS

MFM, LMG, JAM conceived the study. MFM obtained research funding. MFM supervised the data collection and CF extracted the data. SDP provided statistical advice on study design and analyzed the data. MFM drafted the manuscript, and all authors contributed substantially to its revision. MFM takes responsibility for the paper as a whole.

### CONFLICT OF INTEREST SATEMENT

None

## DATA AVAILABILITY

The data underlying this article cannot be shared publicly in order to protect the privacy of individuals represented in the dataset.

## Notes

**Funding:** Dr. Melanie Molina is supported by the National Institute on Drug Abuse (K23DA060993) and the Harold Amos Medical Faculty Development Program. These funders had no role in the design and conduct of the study; collection, management, analysis, and interpretation of the data; preparation, review, or approval of the manuscript; and decision to submit the manuscript for publication. Dr. Melanie Molina had full access to all the data in the study and takes responsibility for the integrity of the data and the accuracy of the data analysis.

### Competing Interest Statement

The authors have declared no competing interest.

### Funding Statement

This study was funded by the National Institute on Drug Abuse (K23DA060993).

### Author Declarations

The University of California, San Francisco Institutional Review Board gave ethical approval for this work.

### Summary of Updates

The measures of engagement with SDOH data have been revised.

