## Appendix A for "Characterizing emergency clinician engagement with social drivers of health data among patients with opioid use disorder"

**Appendix A. Encounters Meeting Opioid Use Disorder Phenotype Criteria, by Criterion**

|  | Count | Proportion^a^ |
| --- | --- | --- |
| ≥1 ICD-10 codes indicating OUD, withdrawal, or overdose in encounter (discharge) diagnoses, problem lists, or past medical history | 3228 | 0.566 |
| Positive urine toxicology results for methadone, fentanyl, or heroin | 1987 | 0.349 |
| ED clinician notes (including triage nursing notes) containing “OUD”, “opioid use disorder”, “opioid use d/o”, “COWS”, “fentanyl”, and/or “heroin” | 1701 | 0.298 |
| Active prescriptions for buprenorphine or methadone preceding the index ED visit encounter | 955 | 0.168 |
| Addiction care team consultation notes with reason for consult containing “heroin”, “opioid”, “opiate”, “Narcan”, and/or “fentanyl” | 562 | 0.099 |
| Reason for visit free text containing “heroin”, “opioid”, “opiate”, and/or “OUD”, “fentanyl” (excluding use together with phrases suggesting therapeutic use or EMS-only administration), “fentanyl use” | 493 | 0.086 |
| ED admission diagnoses containing “heroin”, “opioid”, “opiate”, and/or “OUD”, “fentanyl” (excluding use together with phrases suggesting therapeutic treatment or EMS-only administration), “fentanyl use” and/or mapping to an ICD-10 code for OUD | 36 | 0.006 |

**Abbreviations:** ICD-10, International Classification of Diseases, Tenth Revision; OUD, opioid use disorder; COWS, Clinical Opiate Withdrawal Scale; EMS, emergency medical services; ED, emergency department

^a^Proportions do not add to 1 due to encounters meeting more than one phenotype criteria
